# Pricing Treatments Cost-Effectively When They Have Multiple Indications: Not Just a Simple Threshold Analysis

**DOI:** 10.1101/2023.02.07.23285597

**Authors:** Jeremy D. Goldhaber-Fiebert, Lauren E. Cipriano

**Author notes:** Corresponding Author: Jeremy Goldhaber-Fiebert, Stanford University, Encina Commons, Room 220, 615 Crothers Way, Stanford, CA, 94305, USA.

## Abstract

**Background:** Economic evaluations of treatments increasingly employ price-threshold analyses. When a treatment has multiple indications, standard price-threshold analyses can be overly simplistic. We examine how rules governing indication-specific prices and reimbursement decisions impact value-based price analyses.

**Methods:** We analyze a two-stage game between two players: the therapy’s manufacturer and payer purchasing it for patients. First, the manufacturer selects a price(s) that may be indication-specific. Then, the payer decides whether to provide reimbursement at the offered price(s). We assume known indication-specific demand. The manufacturer seeks to maximize profit, requiring non-negative profit. The payer seeks to maximize total population incremental net monetary benefit and will not pay more than their willingness-to-pay threshold. We consider game variants defined by constraints on the manufacturer’s ability to price and payer’s ability to provide reimbursement differentially by indication.

**Results:** When the manufacturer and payer can both make indication-specific decisions, the problem simplifies to single-indication price-threshold analyses, and the manufacturer captures all the consumer surplus. When the manufacturer is restricted to one price and the payer must make an all-or-nothing reimbursement decision, the selected price is a weighted average of indication-specific threshold prices such that reimbursement of the more valuable indications subsidize reimbursement of the less valuable indications. With a single price and indication-specific coverage decisions, the manufacturer may select a high price and fewer patients receive treatment than in the first-best solution. However, there are also cases when the manufacturer selects a low price resulting in reimbursement for all indications and positive consumer surplus.

**Conclusions:** When multiple indications exist for a given treatment, economic evaluations including price-threshold analyses should carefully consider jurisdiction-specific rules regarding pricing and reimbursement decisions.

## Introduction

Prices of new treatments developed for conditions from cancer to heart disease are rising.^1,2^ For payers and health systems interested in efficiently achieving better health for their populations, price negotiation and value-based pricing are essential. To support such negotiations, evidence in the form of price-threshold analyses is an important feature of economic evaluations, especially those conducted to assess treatments with potentially large budget impacts.

For treatments with a single indication (i.e., disease, condition, or patient sub-population), the threshold analysis is straightforward: the price of the treatment below which it is cost-effective for use for that indication (i.e., at or below the payer’s willingness-to-pay threshold). When there are multiple indications but separate prices can be charged for each, the analysis is again straightforward as it is separable by indication. Such separate threshold analyses will identify higher threshold prices for indications with more per-patient value (i.e., those with higher incremental Net Monetary Benefit at any given treatment price).

However, the price-threshold analysis can be substantially more complicated when a treatment that has multiple indications is considered for reimbursement in systems or situations where it is not feasible or permissible to pay separate indication-specific prices.^3^ In such cases, a single price is required for all indications, raising the question of what single price should be charged.

Similarly, payers may be able to use pre-authorized reimbursement or restricted formularies to enforce indication-specific reimbursement decisions. However, there may also be situations where the financial or political costs of implementing such mechanisms for differentiating indications may be too high or where it may be unethical to differentiate sub-populations, resulting in an all-or-nothing reimbursement decision.

There are multiple real-world examples of situations where a single therapy has multiple indications but where differential pricing and reimbursement by indication may be more or less feasible. These range from a therapy used for the same disease with patients of different characteristics or severity; a therapy used for the same pathogen that causes different diseases across patients groups; or, a therapy used to treat distinct diseases across distinct patient populations. For example, HPV vaccination can prevent a variety of cancers for both females and males. However, given higher rates of cancers, specifically uterine cervical cancer, HPV vaccination costs less per QALY gained for girls than for boys.^4,5,6^ Directly acting antiviral therapies for chronic HCV can arrest the progression of liver fibrosis preventing advanced liver diseases (ALD). However, given that ALD will occur more rapidly and with higher likelihood for chronic HCV-infected 50 year-olds with moderate liver fibrosis than those without liver fibrosis, cost per QALY gained is lower for the former group than the latter.^7^ Chimeric antigen receptor T-cell (CAR-T) therapies are effective in treating a variety of otherwise refractory cancers but can cost $100,000s for a single dose. The cost per QALY gained is lower for CAR-T treatment of relapsed B-cell Acute Lymphoblastic Leukemia in children than for relapsed diffuse large B-cell lymphoma in adults in part because of the larger number of QALYs that can be gained by preventing a cancer death in a child than in an older adult.^8,9^

Whether a payer can and should negotiate separate prices by indication or make separate reimbursement decisions in these examples and other cases is relevant for population health gains, the efficient use of limited healthcare resources, and fairness to patient sub-populations. Our study uses game theory to analyze the optimal pricing and reimbursement decisions for settings in which the pricing and/or reimbursement decisions may be required to be the same across all indications. Further, we evaluate the welfare implications of these restrictions as well as their effects on manufacturer profit and consumer surplus.

## Methods

We analyze a two-stage game between two players: the pharmaceutical manufacturer and the payer who purchases treatment(s) on behalf of patients. In the first stage, the manufacturer selects a price for the treatment that may or may not be indication-specific. In the second stage, the payer decides whether to reimburse patients for the treatment at the offered price(s) for one or more possible treatment indications. We assume that without reimbursement for their indication, patients are not treated. We index the indications with *k* where *k* ∈ {1, 2, …, *K*}). The number of people eligible for treatment per indication is denoted *q*_*k*_. Each player seeks to maximize their respective objective function. We consider scenarios in which we impose constraints on the manufacturer’s ability to price differentially and/or the payer’s ability to provide reimbursement differentially by indication. We compare the optimal outcome of the game sequence in each of these scenarios to the socially optimal outcome without any such constraints.

### Decisions

The game involves a sequence of decisions: pricing decisions made by the manufacturer and then reimbursement decisions made by the payer.

The manufacturer chooses the price for each indication, *p*_*k*_, greater than or equal to their constant marginal cost of production, *c >* 0, such that *p*_*k*_ > *c*. In situations where separate, indication-specific prices are not possible, the manufacturer chooses a single price, i.e., *p*_*k*_ = *p* ∀*k*. For simplicity, we assume that the price is paid as a one-time up-front cost of treatment; but this price can also represent the net present value of treatment costs that would be incurred over time.

The payer then chooses whether or not to reimburse the treatment at the manufacturer’s offered price(s). We denote the payer’s decision with *d*_*k*_ ∈ (0, 1) where *d*_*k*_ = 0 indicates the payer does not, and where *d*_*k*_ = 1 indicates the payer does, reimburse the treatment for indication *k*. For various reasons, the payer may be restricted to only a single decision reimbursing the treatment for all possible indications such that *d*_*k*_ = *d* ∀*k*.

There are three variations of this game that may occur in practice (Table 1). First, both the manufacturer and the payer may be able to make indication-specific decisions. The manufacturer may be able to offer different prices because of very different dosing or different means of delivery, and the payer may similarly be able to easily differentiate how the product is being used on that same basis. Second, the manufacturer is restricted to a single price, and the payer is restricted or uninterested in making an indication-specific decision due to difficulty or cost associated with verifying the specific indication or due to the ethical dimensions of providing differential access. Third, the manufacturer is restricted to a single price, potentially because of a common formulation and dose range, but it may be possible for the payer to use pre-authorized approval to impose different reimbursement decisions by indication. We do not study the fourth possible case, in which the manufacturer may use indication-specific pricing and the payer is limited to a single reimbursement decision. We do not believe this case is realistic as the payer could use the same basis as the manufacturer for making an indication-specific decision. Further, if the same reimbursement decision for all indications is the optimal action, it will be identified as the solution to the more general problem.

**Table 1.**
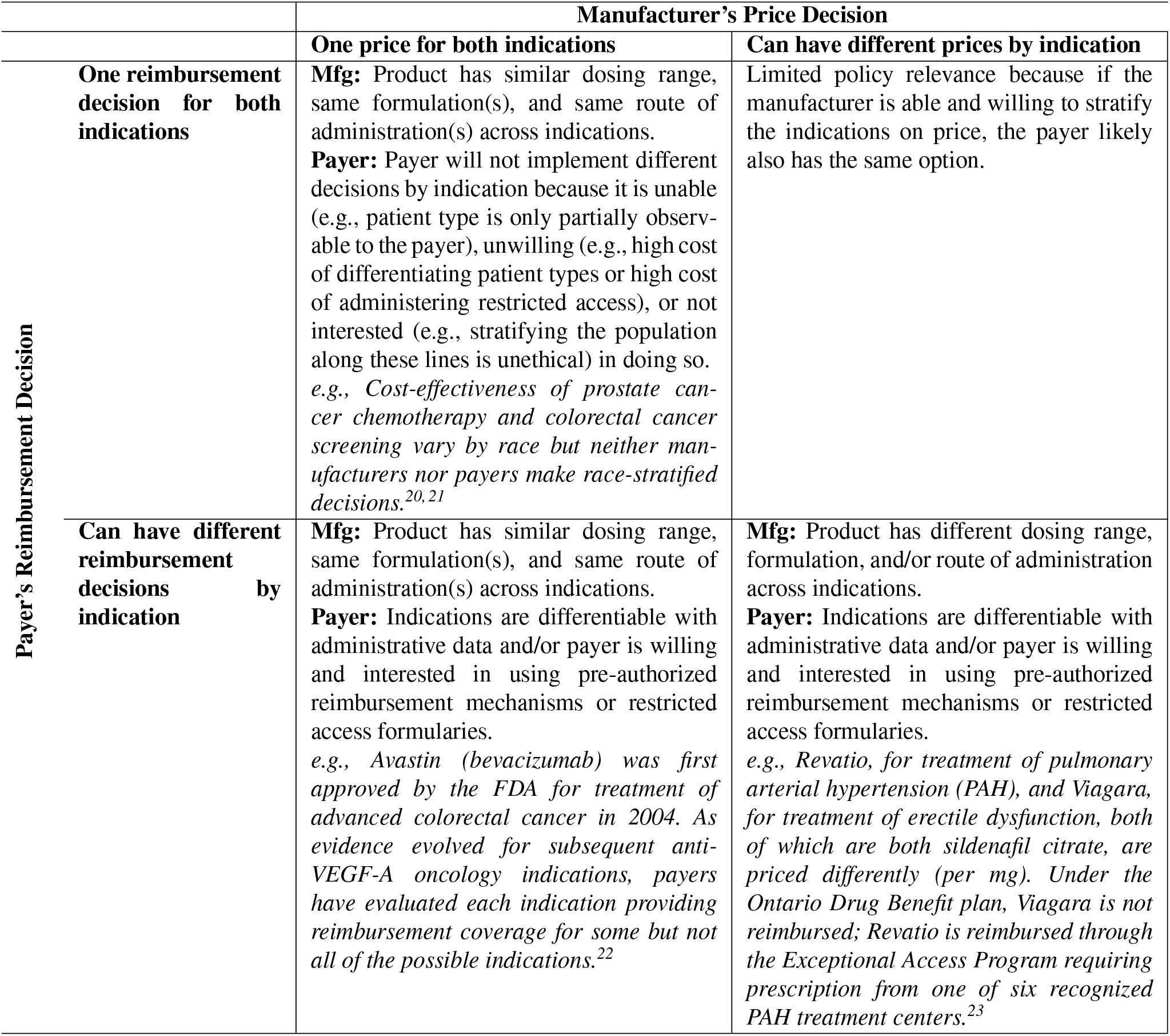
Policy relevant scenarios where the manufacturer may have the ability to price differently by indication and the payer may have the ability to make different decisions by indication.

### Objective Functions

The manufacturer seeks to maximize profit (*π*) where

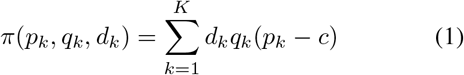

For simplicity, we utilize a linear profit function of the per-patient prices charged for each indication, the quantities of each treatment sold (*q*_*k*_), and the fixed marginal cost of production for the treatment (*c*).

The payer seeks to maximize total population incremental net monetary benefit (*PINMB*). The per-patient incremental net monetary benefit for delivering treatment for indication *k* is defined as

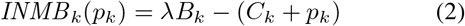

where *λ* is the willingness to pay threshold (WTP) for health benefits; *B*_*k*_ is the per-patient expected discounted lifetime incremental health benefits for indication *k*; and *C*_*k*_ is the expected discounted lifetime per-patient incremental cost excluding the price of treatment *p*_*k*_. The population incremental net monetary benefit for indication *k* depends on the size of the indication-specific patient population (*q*_*k*_) and is then defined as Finally, the total population incremental net monetary benefit for all indications, the payers objective function, is defined as

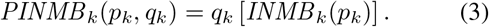

Finally, the total population incremental net monetary benefit for all indications, the payers objective function, is defined as

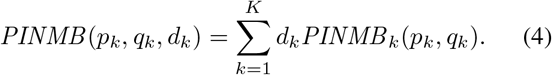

Note that the *PINMB* also represents the consumer surplus, the amount the payer would be willing to pay over and above the amount they actually pay based on the incremental value they place on the treatment (for all indications).

The total social welfare, *W*, can be represented as the sum of the manufacturer’s profit f unction a nd t he payer’s *PINMB* :

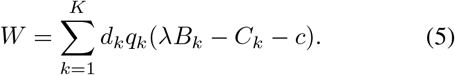

### Participation Constraints

The participation constraints for the manufacturer and the payer respectively are

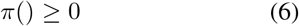

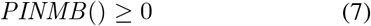

If the participation constraints are not satisfied, then either the manufacturer will opt not to sell the treatment as it is unprofitable to do so or the payer will opt not to reimburse use of the treatment as its overall incremental costs are larger than the value of its overall incremental benefits at the payer’s willingness-to-pay threshold.

### Numerical Analysis

To assist in the visualization of our results, we graphically present a numerical analysis. In this analysis, we assume *q*_1_ = 3000, *q*_2_ = 1500, and *c* = 4. We examine the results of each of the three games across a range of *λB*_*k*_ − *C*_*k*_ values for each of the two indications. We determine the optimal policy regions based on the outcome of each game sequence. We calculate the total welfare, manufacturer profits, and consumer surplus for indications whose per-person values range from − 5 through +30 and determine which game structure is preferred by each agent.

## Results

### Social welfare maximizing policy

We first identify the reimbursement policy decision that will maximize social welfare, *W*, from the societal perspective, where the price paid for treatment above the marginal cost represents a transfer cost between agents in the society. In this analysis we assume that it is possible to make indication-specific reimbursement decisions. Because Equation 5 is linear in the incremental net benefit over and above the marginal production cost, the optimal decision for each indication *k*, is

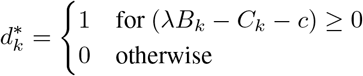

This set of decisions provides patients access to all indications for which there is potential for positive net benefit. We refer to this policy as the “first-best” and we compare other policies resulting from the two-stage game between manufacturers and payers to this policy outcome in later sections.

### Indication-Specific Prices and Reimbursement Decisions

In this section, we first focus on the interesting case where the incremental net benefit of treatment, excluding the cost of treatment, is at least the marginal cost of producing the treatment, i.e., *λB*_*k*_ − *C*_*k*_ ≥ *c*.

When the offered prices can be indication-specific, the range of feasible prices at which both the manufacturer and payer wish to participate can be evaluated separately and independently for each indication. For each indication, feasible price ranges based on participation constraints exist where

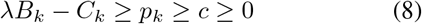

Because the manufacturer’s profit function is linear in each *p*_*k*_, the manufacturer will offer the maximum prices acceptable to the payer. As a result, the offered and accepted prices, 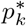, can be found directly using the payer’s participation constraint

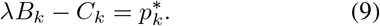

For any indication for which the incremental net benefit of treatment, excluding the cost of treatment, is not at least the marginal cost of production, i.e., *λB*_*k*_ − *C*_*k*_ *< c*, there is no mutually agreeable price at which the payer will provide reimbursement. The manufacturer will not offer a price less than their marginal cost of production, i.e., 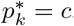, and the highest acceptable price for the payer is below this marginal cost. Thus, for these indications, the payer’s optimal decision will be not to reimburse for that indication (i.e.,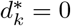).

Ultimately, this situation provides the first-best policy, achieving the highest total welfare, and providing patients access to all indications for which there is potential for positive net benefit, i.e., *λB*_*k*_ − *C*_*k*_ − *c* ≥ 0. However, since the accepted prices will be as high as possible for all reimbursed indications, the manufacturer captures all of the surplus and there is no consumer surplus, represented by the population incremental net monetary benefit,

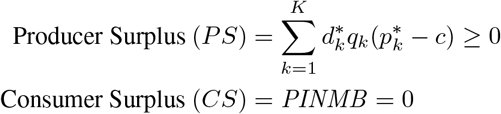

This solution is illustrated when *K* = 2 in Figure 1a and Figures 2a-2c creating four policy regions: (I) in which the manufacturer’s minimum price is greater than the incremental net monetary benefit for both indications; (II and III) in which the manufacturer is able to secure a positive reimbursement decision for only one indication for which it can charge the threshold price of that indication; and, (IV) in which the manufacturer is able to secure a positive reimbursement decision for both indications charging their respective threshold prices, i.e., each 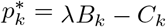.

**Figure 1:**
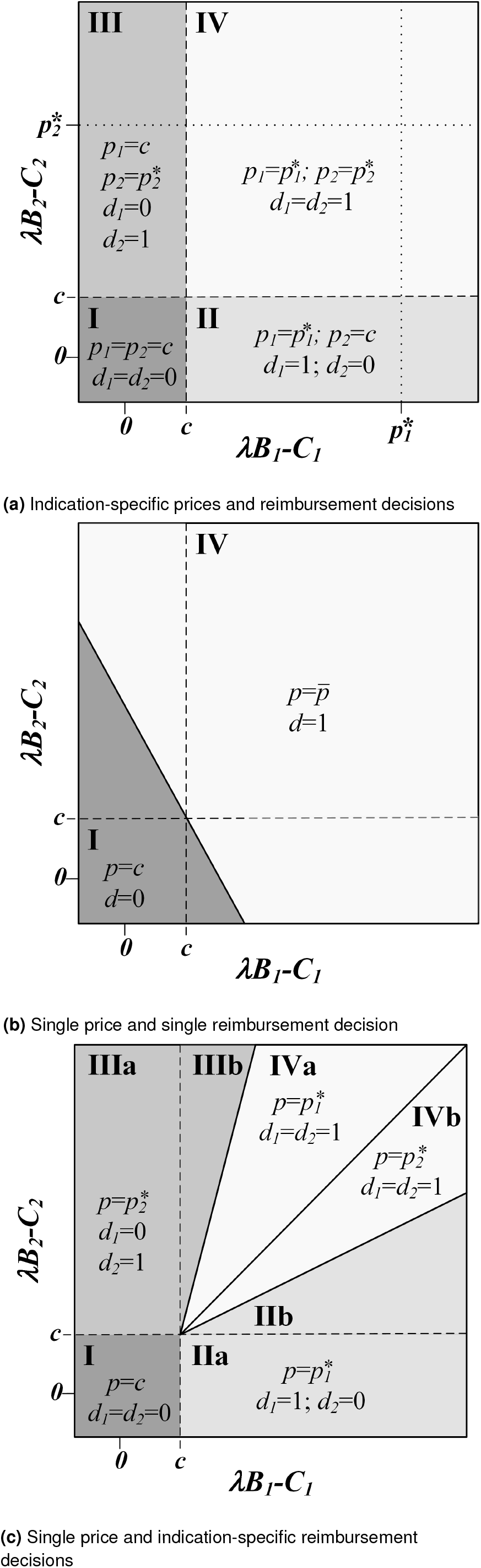
(a) Optimal pricing decisions by the manufacturer, (*p*_1_, *p*_2_), and optimal reimbursement decisions by the payer, (*d*_1_, *d*_2_), at various technology values when it is possible to have indication-specific pricing and indication-specific reimbursement decisions. In region IV, the optimal price, offered and accepted, for each indication is set equal to each indication’s value to society, i.e.,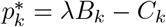. (b) Optimal pricing decision by the manufacturer, *p*, and optimal reimbursement decision by the payer, *d*, at various technology values when both the manufacturer and the payer cannot make indication-specific decisions. In region IV, the optimal price, offered and accepted, is the weighted average of each indication’s value to society, i.e.,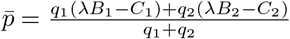. (c) Optimal pricing decision by the manufacturer, *p*, and optimal reimbursement decisions by the payer, (*d*_1_, *d*_2_), at various technology values when the manufacturer is restricted to a single price and the payer is able to make an indication-specific reimbursement decisions. In regions IIb and IIIb, the manufacturer selects a high price that will lead the payer to only reimburse for the higher-value indication, even though at a lower price, it is possible for adoption of the lower-value technology to be cost-effective as well. In regions IVa and IVb, the manufacturer selects a low price that will lead the payer to reimburse for both indications. In region IV, the first best policy is achieved and the payer obtains some consumer surplus associated with the higher-valued indication. The diagonal line between regions IVa and IVb represents all cases where 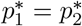.

**Figure 2.**
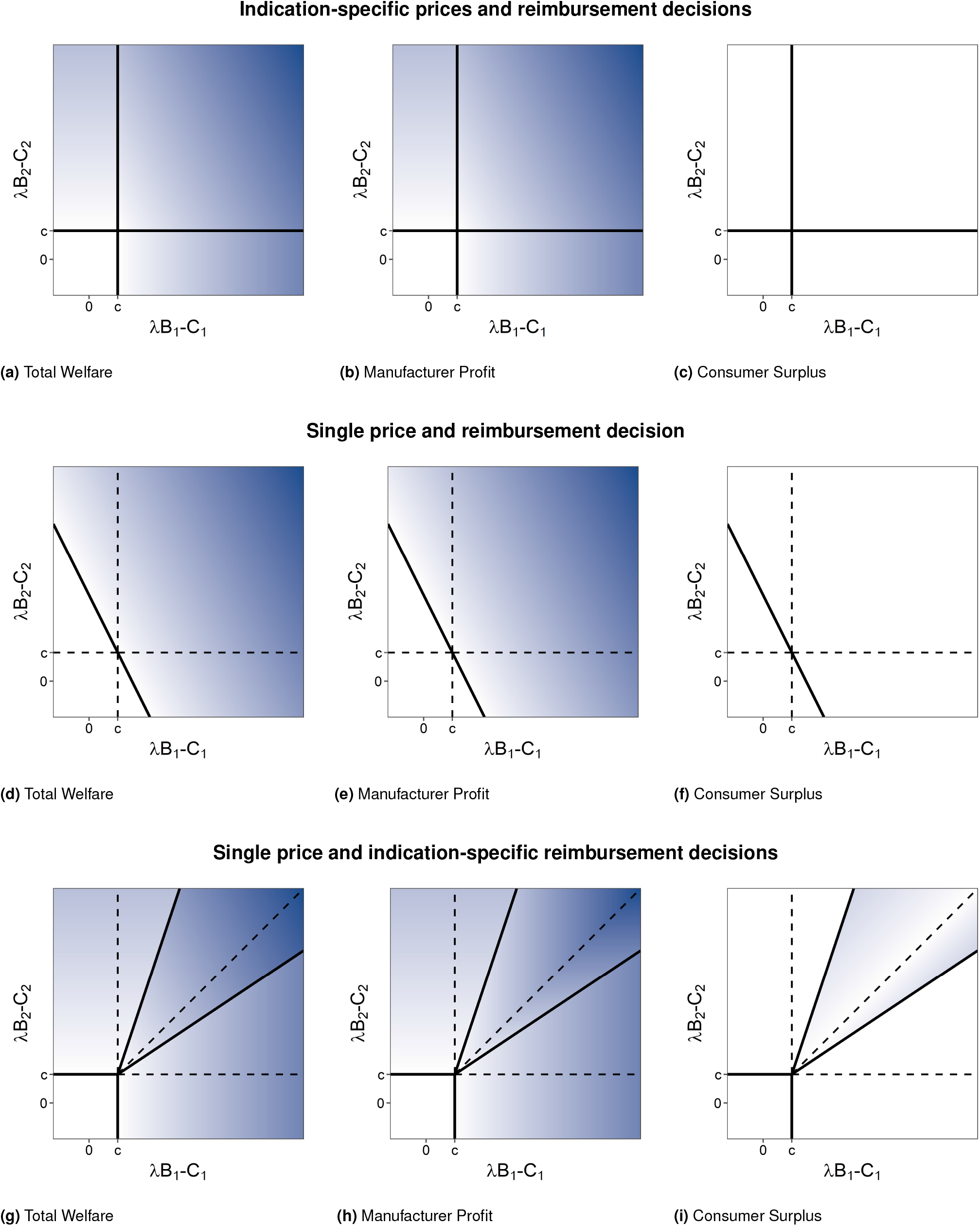
Total Welfare (column 1, panels a, d, and g), Manufacturer Profit (column 2, panels b, e, and h), and Consumer Surplus (column 3, panels c, f, and i) for Indication-specific prices and reimbursement decisions (row 1, panels a-c), Single price and reimbursement decision (row 2, panels d-f), and Single price and indication-specific reimbursement decisions (row 3, panels g-i). Color scale is set such that white is zero and greater intensity of blue represents increasingly positive values.

### Single price and single reimbursement decision for multiple indications

For multiple indications, the single treatment price and single reimbursement decision constraints can be formalized as

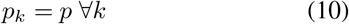

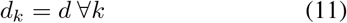

To be feasible from the perspective of the manufacturer, *p* ≥ *c*. The payer’s participation constraint identifies the prices that are acceptable to the payer; setting *d* = 1 and re-arranging the equation reveals the payer’s maximum acceptable price, which we denote 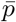:

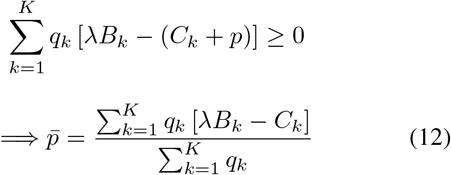

Equation (12) identifies that the maximum price the payer is willing to accept is a weighted average of the value of each indication, with weights determined by the market size for each indication.

As in the previous setting, because the manufacturer’s profit function is linear in *p*, the manufacturer offers the payer’s maximum acceptable price, 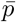, when that price is greater than *c*. However, when one or more indication has an incremental net benefit, excluding the cost of treatment, that is less than the marginal cost of production, i.e., *λB*_*k*_ − *C*_*k*_ < *c*, it is possible that this weighted average will identify the payer’s maximum acceptable price is less than *c*. When this happens, the manufacturer will offer a price equal to its marginal cost, *c*, and the payer will decide not to reimburse for any indication.

Whenever there is a mutually acceptable price, because it is always the payer’s maximum acceptable price, the manufacturer captures all of the surplus and there is no consumer surplus.

This solution is illustrated when *K* = 2 in Figure 1b and Figures 2d-2f creating two policy regions: (I) in which the manufacturer’s minimum price is greater than the payer’s maximum acceptable price leading to a no coverage decision; and, (IV) in which the manufacturer is able to secure a positive coverage decision for both indications by offering the price 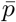 that is the weighted average of each indication’s incremental net benefit excluding the cost of treatment. Note that when one of the two indications has a incremental net benefit, excluding the cost of treatment, that is less than the marginal cost of production, either policy outcome may occur depending on the relative contribution and relative size of each indication. The line separating the two policy regions is identified by solving for the threshold at which the manufacturer will obtain zero profit, 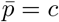, at the payer’s maximum acceptable price:

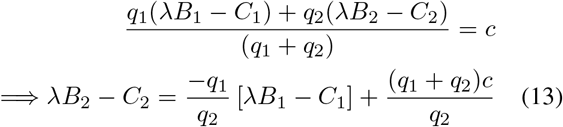

Comparing this setting to that of indication-specific prices and reimbursement decisions, the additional constraints of a single price and a single reimbursement decision affect outcomes for both the manufacturer and the payer (Figures 3a-3c). Regions II and III from Figure 1a are now each split into two parts. One of these parts joins region IV, in which the payer reimburses both indications, providing patients access to the less valuable indication, decreasing social welfare, and decreasing the profits of the manufacturer even as it increases the units sold. The second part joins region I, in which the payer now decides not to reimburse for either indication, removing patient access to the more valuable indication and thereby decreasing social welfare and manufacturer profits. In both altered regions, the payer is not better or worse off as they do not obtain any consumer surplus in either situation, but the total social welfare is reduced relative to that obtained in the corresponding parts of regions II and III of Figure 1a.

**Figure 3.**
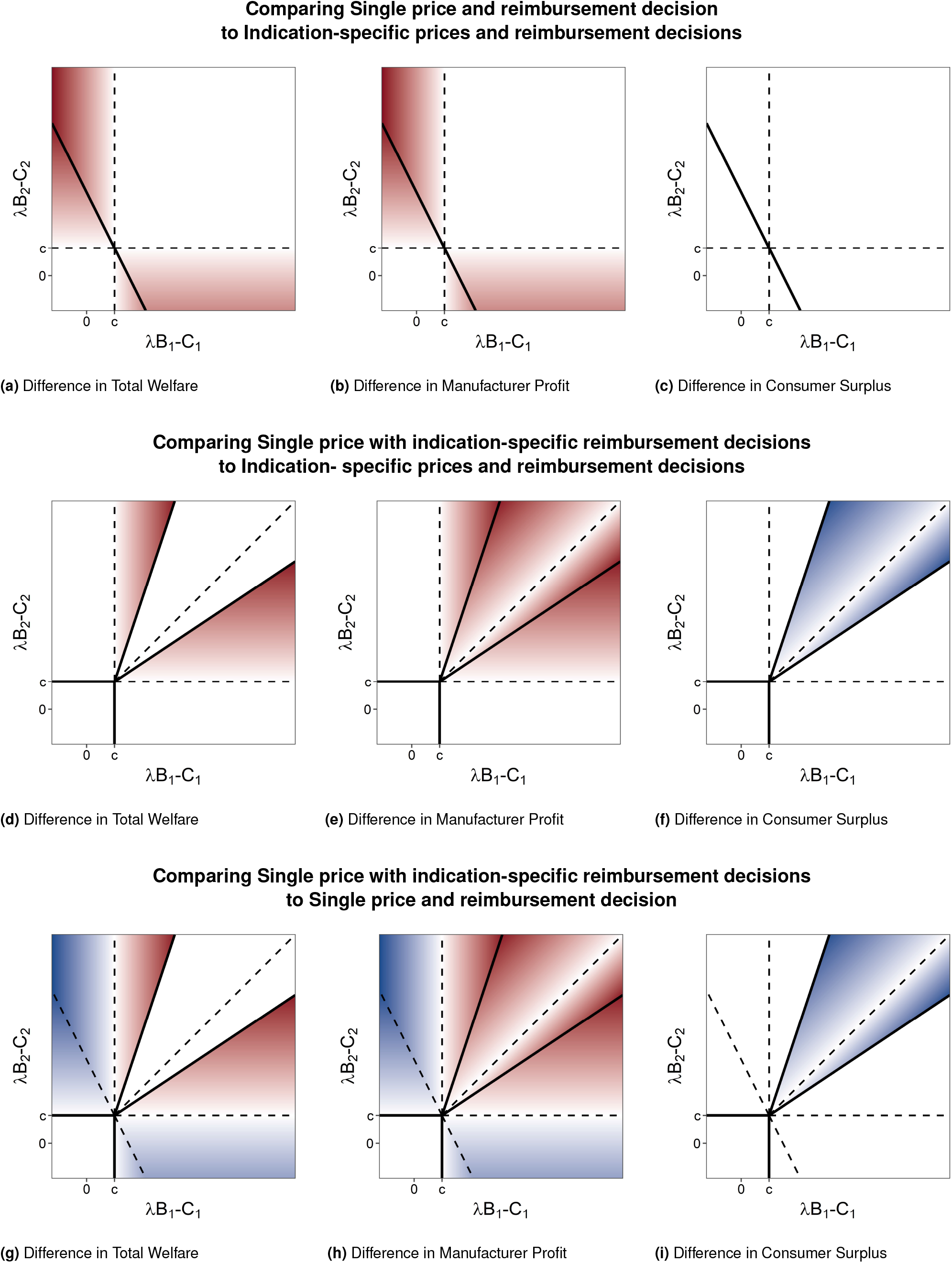
Differences in Total Welfare (column 1), Manufacturer Profit (column 2), and Consumer Surplus (column 3) comparing games with single and indication-specific pricing and/or reimbursement decisions. Color scale is set such that white is zero, greater intensity of blue represents increasingly positive values, and greater intensity of red represents increasingly negative values.

### Single price with indication-specific reimbursement decisions

We now focus on the situation in which there is a single price for all indications but indication-specific reimbursement decisions. In this section, it is useful to rank potential indications based on their incremental net monetary benefit excluding the cost of treatment, i.e., *λB*_*k*_ − *C*_*k*_. So, for each indication, we denote the ranked value of the indication using *k*^(1)^ for the highest valued indication, *k*^(2)^ for the second highest valued indication, etc. The *n* ≤ *K* indications whose values exceed the marginal cost of production can then be written in sequence

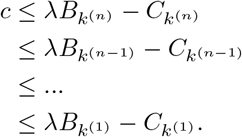

With a ranked list of the treatment indications, it is clear to see that the payer has *K* + 1 possible policy decisions each representing the selection of a threshold above which all indications are reimbursed and none below it. Similarly, in the selection of a single price, the manufacturer’s decision alternatives can be reduced to the set of maximum acceptable prices that would lead to each of those *K* + 1 decisions. For example, the maximum acceptable price that would result in a positive reimbursement decision for all indications equal to or greater than the value of indication *k*^(*x*)^ is 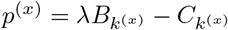. There is no price higher than the threshold price for *k*^(*x*)^ for which the payer will decide to reimburse for this indication.

The manufacturer may prefer to select a price lower than the threshold price for indication *k*^(1)^, when doing so increases volume of sales commensurate with the lost revenue associated with the lower price. When this occurs, the payer is able to absorb some of the surplus. Specifically, when the manufacturer selects the *x*^*th*^ highest valued indication’s threshold price, the payer will decide to reimburse for interventions *k*^(*x*)^, *k*^(*x*−1)^, *k*^(*x*−2)^, …, *k*^(2)^, and *k*^(1)^, and the producer surplus is

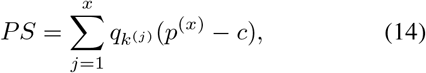

and the consumer surplus is

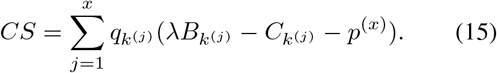

This solution is illustrated when K=2 in Figure 1c and Figures 2g-2i with four policy regions: (I) in which the manufacturer’s minimum price is greater than the incremental net monetary benefit for both indications; (II and III) in which the manufacturer is able to secure a positive reimbursement decision for only one indication for which it can charge the threshold price of that indication; and, (IV) in which the manufacturer is able to secure a positive reimbursement decision for both indications. Region IV is further divided into two sections, each in which the price selected to secure the ‘reimburse all’ decision is the threshold price for the lower-valued indication.

Comparing this setting to that of indication-specific prices and reimbursement decisions, there are several differences for the manufacturer and the payer (Figures 3d-3f). First, regions II and III, in which only one indication is reimbursed are larger. In the regions of Figure 1c labelled IIb and IIIb, the payer selects the threshold price of the higher-valued indication, forgoing market access to the lower-valued indication, because the incremental sales that would come from the lower-valued indication do not offset the lost revenue from the reduced price charged for the higher-valued indication. In these regions, the outcome of the game sequence is no longer the first-best solution for society (Figure 3d). Compared to the first-best solution, total welfare is lower because there is no reimbursement for an indication that has the potential to provide positive net benefit (at some price satisfying the participation constraints). Compared to the setting in which there are both indication-specific prices and decisions, manufacturer profits are reduced. In both settings, the payer receives no consumer surplus in these regions.

In the regions labelled IVa and IVb in Figure 1c, the manufacturer offers the threshold price associated with the lower-valued indication maximizing profits through a larger total volume of sales. In these regions, the policy outcome of the game sequence is the first-best solution for society. Compared to the setting in which there are both indication-specific prices and decisions, manufacturer profits are reduced with the surplus now shared between the manufacturer and the payer (Figures 3e and 3f).

The line separating regions IIb and IVb (and similarly regions IIIb and IVa) is identified by solving for the manufacturer’s indifference point where they make the same profit from selling only the higher-valued indication at the higher price as they do selling both indications at the lower-valued indication’s threshold price.

Line separating region IIb and IVb:

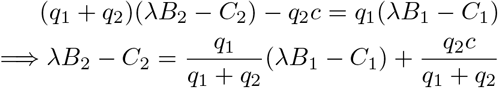

Line separating region IIIb and IVa:

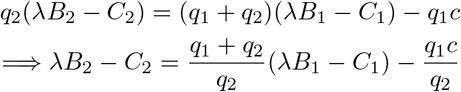

These equations reveal the importance of the relative size of each indication’s treatment population. If the higher-valued indication has a relatively large treatment population, the manufacturer will offer the higher price, expanding region IIb (or IIIb). If the higher-valued indication has a relatively small treatment population or difference in value between the two indications is relatively small, the manufacturer is willing to forgo some otherwise obtainable revenue for the higher-value indication in favor of sales of both indications, enlarging regions IVa (or IVb).

### Comparing outcomes with a single vs. indication-specific reimbursement decisions

Comparing outcomes with a single price and indication-specific reimbursement decisions to that of a single decision for all indications provides insight into which game is preferred by manufacturers and payers (Figures 3h-3i). In regions labelled IIa and IIIa in Figure 1c, indication-specific decisions are preferred by the manufacturer because otherwise the lower-valued indication negatively impacts the weighted-average single price, lowering manufacturer profits. The payer has the same consumer surplus in both settings, but when there are indication-specific decisions, all indications for which there is positive net monetary benefit, excluding the cost of treatment, are reimbursed, allowing patients from more indications access to effective treatments and increasing overall welfare.

When both indications have positive incremental net monetary benefit, excluding the cost of treatment, the manufacturer always prefers a single decision. In regions labelled IIb and IIIb on Figure 1c this is because the manufacturer optimizes their profit by choosing the higher-value indication’s threshold price, forgoing market access to the lower-value indications sales and, overall, having less profit than would occur with a single decision. In regions labelled IVa and IVb on Figure 1c, the manufacturer prefers a single decision because the manufacturer maximizes their profit by choosing the lower-valued indication’s threshold price, and while this results in market access for both products, it does so at a lower price than the weighted average price that occurs when the payer must make a single decision.

In regions IIb and IIIb, the payer has the same consumer surplus (none) regardless of whether they make indication-specific decisions or not. However, in these regions, social welfare is higher because both indications are ultimately reimbursed when there is a single decision. In contrast, in regions IVa and IVb, the payer strictly prefers indication-specific decisions. In these regions, the same policy decision is made in both settings, leading to the same overall welfare. However, in the setting of indication-specific decisions, the manufacturer selects a lower price in order to ensure that both indications are given market access.

## Discussion

Our study analyzes three different two-stage pricing/reimbursement games, in which the pricing and/or reimbursement decisions may be required to be the same across all indications. Our study shows that value-based reimbursement decisions are substantially more complex than a simple price-threshold analysis when a given treatment has more than one indication and if either indication-specific pricing or indication-specific reimbursement decisions are disallowed. The implication is that, when a treatment has more than one indication, analysts should be cautious in claiming that because a treatment becomes cost-effective for a specific indication at an identified price-threshold, the manufacturer should, therefore, alter the price accordingly and that likewise, the payer should not reimburse the treatment for that indication unless this is done. Instead, caution and nuance are warranted when separate indication-specific prices and indication-specific reimbursement decisions are not permitted.

Our analysis shows that the socially optimal outcome, in which patients have access to all treatments for which the incremental net monetary benefit, excluding the marginal cost of production, can only be guaranteed when every treatment, or treatment-indication pair, is priced commensurate with its own incremental net monetary benefit. Practical, logistical, or ethical considerations can effectively impose limits on whether a manufacturer or a payer can make indication-specific decisions (Table 1). In situations where a manufacturer is effectively restricted to a single price because its product has the same formulation, a similar dosing range, and the same routes of administration across indications, payers may be able to choose between making a single reimbursement decision or indication-specific decisions using mechanisms like pre-authorized reimbursement schemes or restricted formularies. However, there may also be situations where the financial or political costs of implementing such mechanisms for differentiating indications may be too high or where it may be unethical to differentiate sub-populations.

In the standard, single-indication value-based pricing analysis, the price is set, irrespective of the size of the patient population, such that the manufacturer absorbs all of the gains, and there is no consumer surplus. The extension of this to separate, indication-specific pricing and reimbursement decisions yields the same results for each indication. However, when indication-specific pricing or reimbursement decisions are not possible, our study shows that using a price representing the average value across indications, weighted by the relative size of each indication’s patient population, can achieve the first-best reimbursement policy when all indications contribute positive net incremental benefit excluding the cost of treatment.

However, the feasibility of successful implementation such a weighted-average single price is potentially vulnerable to asymmetries of information between the manufacturer and the payer, to uncertainty in the treatment effectiveness as well as the size of the treatment population, to the possibility of new entrants into the marketplace, or to gaming by the manufacturer with respect to strategic sequential introduction of indications.

One key challenge is in the estimation of the quantity demanded for each indication, *q*_*k*_. Often new product uptake is highly uncertain and the quantity sold for each indication can depend on the relative effectiveness of current treatments.^10^ The quantity sold for each indication can also be manipulated by the manufacturer through differential marketing effort^11^ or by the payer through investment in adoption or quality improvement programs.^12,13^ Because, in such cases, higher-valued indications function as sponsors, lending their benefits to subsidize reimbursement for lower-value indications, the optimal price to offer and accept depends directly on actual indication-specific demand. Ultimately, there are risks to both the manufacturer and payer if the realized demand differs from what was originally estimated when determining the single price.

A single price and reimbursement decision scheme is also susceptible to innovation affecting the indication mix over time. Changes to the indication mix can happen in numerous ways. First, through additional study, evidence may emerge that the treatment is less effective for one or more indications than initially estimated, or that there is a previously unknown lower-value indication. In some situations it may be possible for a manufacturer to strategically lead with market introduction of higher-valued indications, garnering a high price, and only then introduce evidence supporting use of the product in lower-valued indications. Second, a new product can enter the market replacing the optimal treatment or influencing the incremental value for one or more indication. Because pricing in this setting is linked across indications, the entrance of a new treatment relevant to a subset of the indications in the original game can disrupt the price it determined. This disruption can have surprising and unintended effects such as leading to the loss of reimbursement for other (lower-value) indications that perhaps have no direct relationship to the new treatment.

For example, bevacizumab was first approved for the treatment of advanced colorectal cancer in 2004 with new indications identified over time,^14^ which could have destabilized other prices or market access in a strict single price and single reimbursement decision situation. Regardless of how the indication mix changes over time, the payer may end up overpaying for the remaining indications for which the treatment remains in use unless there is an ongoing, value-based market access and price negotiation. However, reimbursement decisions are notoriously difficult to reverse, giving payers little power to effectively renegotiate prices.^15^ As a result, even when our analysis indicates payers may be indifferent between indication-specific and single reimbursement decisions scenarios, other practical considerations not captured in our model may lead payers to strictly prefer indication-specific decisions.

While our study analyzes each of the three price/reimbursement games separately, in principle, decisions by either party or negotiations between them could determine which of these games to engage in, effectively generating a unified three- or four-stage game or a series of sequential games. For example, the manufacturer may choose indication-specific pricing and present the decision problems as unrelated in an effort to secure a higher price for a higher-valued indication (e.g., bevacizumab and ranibizumab for neovascular macular degeneration^16,17^). Similarly, manufacturers may present a single decision problem when the payer is better off making indication-specific decisions in order to select only indications with positive incremental net benefit (e.g., tumour agnostic chemotherapies^18^). Ultimately, when the manufacturer is restricted to a single price, there is no guarantee of achieving the socially optimal policy. When the payer can make indication-specific reimbursement decisions, we have shown that the manufacturer may select a price to maximize profit that will lead to some potentially good-value indications being excluded from reimbursement. In contrast, when the indications have relatively similar value, the selection of a lower price aimed at ensuring a positive reimbursement decision for all indications leads to the first-best policy outcome, albeit at a lower profit for the manufacturer as the payer accrues some consumer surplus. In this latter case, it is easy to see that the manufacturer may prefer one of the two-stage games over the others.

In addition to the considerations discussed in the previous paragraphs, our analysis has a number of other limitations. It makes a number of simplifying assumptions which avoid complicating the mathematical exposition but do not impact the implications of our findings. First, we only consider linear profit functions. Second, we assume the marginal cost of production is the same for all indications. Third, we assume that all parameters in our analysis are known and constant. Uncertainty in the effectiveness of one or more indication may exceed the payers willingness to accept risk; more information may be warranted.^19^ This is another setting in which indication-specific decisions may be preferred by both manufacturers and payers, as to not have uncertainty in some indications affect the timeline for access to treatment for other indications.

Our analysis and findings apply to a broad range of decisions beyond those regarding drugs or other treatments that are effective across numerous diseases. It also applies to cases where indications are not separate diseases but rather subgroups of individuals defined by their risk level or severity of illness within a given disease or condition. For example, treatment with a drug for stage 2 hypertension may yield greater *INMB* than the same treatment for stage 1 hypertension. Yet, if the number of patients with stage 2 hypertension is relatively small, total profits for the manufacturer may be substantially higher if there is a single reimbursement decision, even if the single price charged for both indications is lower than what could have been charged for stage 2 hypertension. While differentiating separate diseases as indications is relatively straightforward, how to divide subgroups of patients into separate indications is far more nuanced with potentially diverging strategic interests on the part of the manufacturer and payer on when and how to do this.

## Conclusions

For economic evaluations, price-threshold analyses are simple only when treatments have a single indication or where pricing and reimbursement by indication are permitted and feasible. However, this is infeasible in many jurisdictions and settings. Furthermore, many treatments have multiple indications in terms of providing benefits to people with different diseases or in terms of providing benefits to several subgroups of patients with the same disease whose expected magnitude of benefit and/or costs differ. The result is that at any given price, the treatment has a different incremental cost-effectiveness ratio for each of these groups. In such situations, the inability to price by indication or to reimburse by indication makes the analysis much more complex and nuanced. Hence, analysts should be cautious when conducting economic evaluations that include treatments with multiple indications. Likewise, policymakers should be cautious in implementing rules that link pricing or reimbursement decisions across indications as they can have unintended, negative implications for social welfare and health equity.

## Data Availability

All data produced in the present study are available upon reasonable request to the authors

